# Integration of polygenic and gut metagenomic risk prediction for common diseases

**DOI:** 10.1101/2023.07.30.23293396

**Authors:** Yang Liu, Scott Ritchie, Shu Mei Teo, Matti Olavi Ruuskanen, Oleg Kambur, Qiyun Zhu, Jon Sanders, Yoshiki Vazquez-Baeza, Karin Verspoor, Pekka Jousilahti, Leo Lahti, Teemu Niiranen, Veikko Salomaa, Aki Havulinna, Rob Knight, Guillaume Méric, Michael Inouye

## Abstract

Multi-omics has opened new avenues for non-invasive risk profiling and early detection of complex diseases. Both polygenic risk scores (PRSs) and the human microbiome have shown promise in improving risk assessment of various common diseases. Here, in a prospective population-based cohort (FINRISK 2002; n=5,676) with ∼18 years of e-health record follow-up, we assess the incremental and combined value of PRSs and gut metagenomic sequencing as compared to conventional risk factors for predicting incident coronary artery disease (CAD), type 2 diabetes (T2D), Alzheimer’s disease (AD) and prostate cancer. We found that PRSs improved predictive capacity over conventional risk factors for all diseases (ΔC-indices between 0.010 – 0.027). In sex-stratified analyses, gut metagenomics improved predictive capacity over baseline age for CAD, T2D and prostate cancer; however, improvement over all conventional risk factors was only observed for T2D (ΔC-index 0.004) and prostate cancer (ΔC-index 0.005). Integrated risk models of PRSs, gut metagenomic scores and conventional risk factors achieved the highest predictive performance for all diseases studied as compared to models based on conventional risk factors alone. We make our integrated risk models available for the wider research community. This study demonstrates that integrated PRS and gut metagenomic risk models improve the predictive value over conventional risk factors for common chronic diseases.

## Introduction

Multi-omic technologies have uncovered new biomarkers for various common diseases, including cardiovascular disease, diabetes, liver disease, dementia and cancer[1–6]. While conventional risk prediction typically relies on demographic (e.g. age or sex), anthropomorphic (e.g. body mass index), lifestyle factors and disease-specific clinical laboratory measurements (e.g. blood pressure, non-HDL cholesterol, mammographic density, creatinine, HbA1c), the recent emergence of multi-omics means that it is now possible to measure and integrate whole classes of biomolecular and cellular factors for the purposes of building multi-omic risk scores.

Polygenic risk scores (PRSs), a quantitative measure of genetic predisposition for a phenotype, have demonstrated validity and potential clinical utility in risk prediction for various common diseases[7–10], for example in cardiovascular disease[11–14], cancers[15, 16], diabetes mellitus[17–19] and ankylosing spondylitis[20]. Given the potential of a genome-wide genotyping array as a one-time, relatively inexpensive assay from which hundreds of PRSs can be calculated, PRSs are being assessed in clinical studies for healthcare systems around the world[9, 11, 21].

The gut microbiota (the collection of microorganisms inhabiting the human gastrointestinal tract) has also been shown to have a role in many common diseases[22–24]. Gut microbial signatures have been associated with incident diseases in the general population, such as type 2 diabetes, liver and respiratory diseases[4, 25–28], suggesting the potential of the gut microbiome in disease risk prediction. Notably, while genome-wide association studies have revealed the human genetic basis of the gut microbiome[29–31], it is apparent that the heritability of the gut microbiome is relatively low[32, 33].

Given that they are based on robust scalable technologies, use non-invasive sampling and have been applied in numerous disease risk prediction studies, PRSs and the gut microbiome comprise promising components of potential future multi-omic risk prediction[34, 35]. In this study, we investigate the predictive capacity of PRSs, gut microbial composition and conventional risk factors for multiple common diseases. We focus on diseases for which there is prior evidence of substantial predictive capacity for PRSs and the human gut microbiome, i.e. coronary artery disease (CAD)[12, 36], type 2 diabetes (T2D)[26, 37], Alzheimer’s disease (AD)[38, 39] and prostate cancer[40, 41]. We utilised the population-based multi-omic FINRISK 2002 cohort[42] to assess the individual and combined performance of PRSs, gut microbiome scores and conventional risk factors to incident disease. Finally, we generated and validated multi-omic predictive models for each disease and make these available to the research community.

## Results

For those in FINRISK 2002 with imputed genotypes and gut metagenomic sequencing, there were 333 incident CAD, 579 T2D, 273 AD and 141 prostate cancer cases over a median follow-up of 17.8 years through electronic health records (EHRs). Characteristics of the study sample of FINRISK 2002 cohort for each disease are given in **Table 1**. For CAD, T2D and AD, baseline clinical risk factors were significantly different between incident cases and non-cases with the exception of smoking for T2D, and sex, diastolic blood pressure and HDL for AD. We detected significant differences between case and non-case groups in baseline age and smoking for prostate cancer.

**Table 1.** Characteristics of participant risk factors for the diseases studied.

|  | Cases | Non-cases | P value |
| --- | --- | --- | --- |
| **CAD** | n=333 | n=4760 |  |
| Male, n (%) | 225 (67.57) | 2015 (42.33) | $3.62 \times 10^{-19}$ |
| Baseline age (years) | $56.81 \pm 9.74$ | $47.55 \pm 12.40$ | $4.58 \times 10^{-39}$ |
| BMI ($\text{kg/m}^2$) | $27.91 \pm 3.96$ | $26.46 \pm 4.24$ | $4.27 \times 10^{-11}$ |
| Systolic BP (mmHg) | $144.90 \pm 20.07$ | $134.10 \pm 19.36$ | $3.36 \times 10^{-23}$ |
| Total cholesterol (mmol/L) | $6.02 \pm 1.09$ | $5.58 \pm 1.05$ | $9.57 \times 10^{-13}$ |
| HDL (mmol/L) | $1.37 \pm 0.39$ | $1.53 \pm 0.41$ | $1.84 \times 10^{-14}$ |
| Smoking, n (%) | 106 (31.83) | 1165 (24.47) | $3.87 \times 10^{-3}$ |
| Exercise, n (%) | 52 (15.62) | 1182 (24.83) | $9.03 \times 10^{-5}$ |
| Prevalent diabetes, n (%) | 26 (7.81) | 137 (2.88) | $1.56 \times 10^{-5}$ |
| Family history, n (%) | 130 (39.04) | 1142 (23.99) | $4.25 \times 10^{-9}$ |
| **T2D** | n=579 | n=4718 |  |
| Male, n (%) | 306 (52.85) | 2114 (44.81) | $2.84 \times 10^{-4}$ |
| Baseline age (years) | $53.26 \pm 10.57$ | $48.37 \pm 12.89$ | $1.14 \times 10^{-18}$ |
| BMI ($\text{kg/m}^2$) | $29.98 \pm 4.18$ | $26.13 \pm 3.99$ | $1.27 \times 10^{-88}$ |
| Systolic BP (mmHg) | $142.67 \pm 20.81$ | $134.50 \pm 19.65$ | $4.67 \times 10^{-21}$ |
| Total cholesterol (mmol/L) | $5.84 \pm 1.20$ | $5.58 \pm 1.04$ | $2.43 \times 10^{-6}$ |
| HDL (mmol/L) | $1.35 \pm 0.35$ | $1.54 \pm 0.41$ | $9.72 \times 10^{-32}$ |
| Triglyceride (mmol/L) | $1.91 \pm 1.29$ | $1.32 \pm 0.83$ | $8.41 \times 10^{-6}$ |
| Smoking, n (%) | 160 (27.63) | 1155 (24.48) | 0.103 |
| Exercise, n (%) | 82 (14.16) | 1168 (24.76) | $3.80 \times 10^{-9}$ |
| Family history, n (%) | 251 (43.35) | 1159 (24.57) | $2.57 \times 10^{-20}$ |
| **AD** | n=273 | n=5074 |  |
| Male, n (%) | 128 (46.89) | 2349 (46.29) | 0.852 |
| Baseline age (years) | $64.29 \pm 6.52$ | $48.21 \pm 12.46$ | $1.07 \times 10^{-93}$ |
| BMI ($\text{kg/m}^2$) | $28.08 \pm 4.05$ | $26.59 \pm 4.24$ | $1.38 \times 10^{-9}$ |
| Systolic BP (mmHg) | $144.82 \pm 20.59$ | $135.01 \pm 19.90$ | $5.60 \times 10^{-16}$ |
| Diastolic BP (mmHg) | $79.63 \pm 10.08$ | $79.14 \pm 11.17$ | 0.489 |
| Total cholesterol (mmol/L) | $5.84 \pm 1.12$ | $5.57 \pm 1.05$ | $1.07 \times 10^{-4}$ |
| HDL (mmol/L) | $1.50 \pm 0.45$ | $1.51 \pm 0.41$ | 0.304 |
| Alcohol consumption (g/week) | $62.63 \pm 138.15$ | $82.76 \pm 123.58$ | $1.77 \times 10^{-8}$ |
| Smoking, n (%) | 46 (16.85) | 1279 (25.21) | $1.50 \times 10^{-3}$ |
| Exercise, n (%) | 44 (16.12) | 1219 (24.02) | $2.62 \times 10^{-3}$ |
| Prevalent T2D, n (%) | 18 (6.59) | 128 (2.52) | $4.03 \times 10^{-4}$ |
| Prevalent stroke, n (%) | 13 (4.76) | 100 (1.97) | $7.20 \times 10^{-3}$ |
| Prevalent psychiatric disorders, n (%) | 12 (4.40) | 121 (2.38) | 0.045 |
| **Prostate cancer** | **n=141** | **n=2323** |  |
| Baseline age (years) | 59.79 ± 7.66 | 49.39 ± 12.62 | 1.79×10<sup>-22</sup> |
| BMI (kg/m<sup>2</sup>) | 27.45 ± 3.03 | 27.07 ± 3.81 | 0.086 |
| Alcohol consumption (g/week) | 113.70 ± 147.06 | 123.40 ± 152.37 | 0.819 |
| Smoking, n (%) | 23 (16.31) | 716 (30.82) | 1.97×10<sup>-4</sup> |
| Exercise, n (%) | 34 (24.11) | 607 (26.13) | 0.693 |
| Family history, n (%) | 62 (43.97) | 794 (34.18) | 0.022 |

Numerical variables are shown as mean ± SD. Categorical variables are shown as number of individuals and percentage of their respective disease status group. P values of Mann-Whitney U test and Fisher's exact test are reported for numerical and categorical variables, respectively.

### Polygenic risk scores and conventional risk factors

Previously validated PRSs for CAD[12] (PGS000018), T2D[37] (PGS000036), AD[38] (PGS000334),and prostate cancer[40] (PGS000662) were obtained from the Polygenic Score Catalog[43] (**Methods**). Cox regression models were used to assess the predictive performance of PRS and disease-specific conventional risk factors for incident diseases.

We first assessed prediction performance of PRS and conventional risk factors (**Methods**) individually for their respective incident diseases (**Figure 1**). In sex-stratified (except for prostate cancer) Cox models of individual risk factors for incident CAD, AD and prostate cancer, baseline age had the highest C-statistic (0.719, 95% confidence interval (CI) 0.695-0.743; 0.880, 95% CI 0.864- 0.895; and 0.769, 95% CI 0.739-0.798, respectively). For CAD and AD, systolic blood pressure was the second strongest individual factor by C-statistic (0.649, 95% CI 0.619-0.679 and 0.656, 95% CI 0.623-0.688, respectively), followed by comparable C-statistics for PRS (0.626, 95% CI 0.595-0.656 and 0.650, 95% CI 0.616-0.684, respectively). For incident prostate cancer, PRS was stronger than other individual conventional risk factors except baseline age with C-statistic of 0.641 (95% CI 0.593- 0.690). For incident T2D, BMI had the strongest C-statistic (0.745, 95% CI 0.726-0.764) and PRS had C-statistic of 0.612 (95% CI 0.589-0.636), similar to the other conventional risk factors. PRS alone achieved higher C-statistic than family history for all diseases where this was available, including CAD, T2D and prostate cancer.

**Figure 1.**
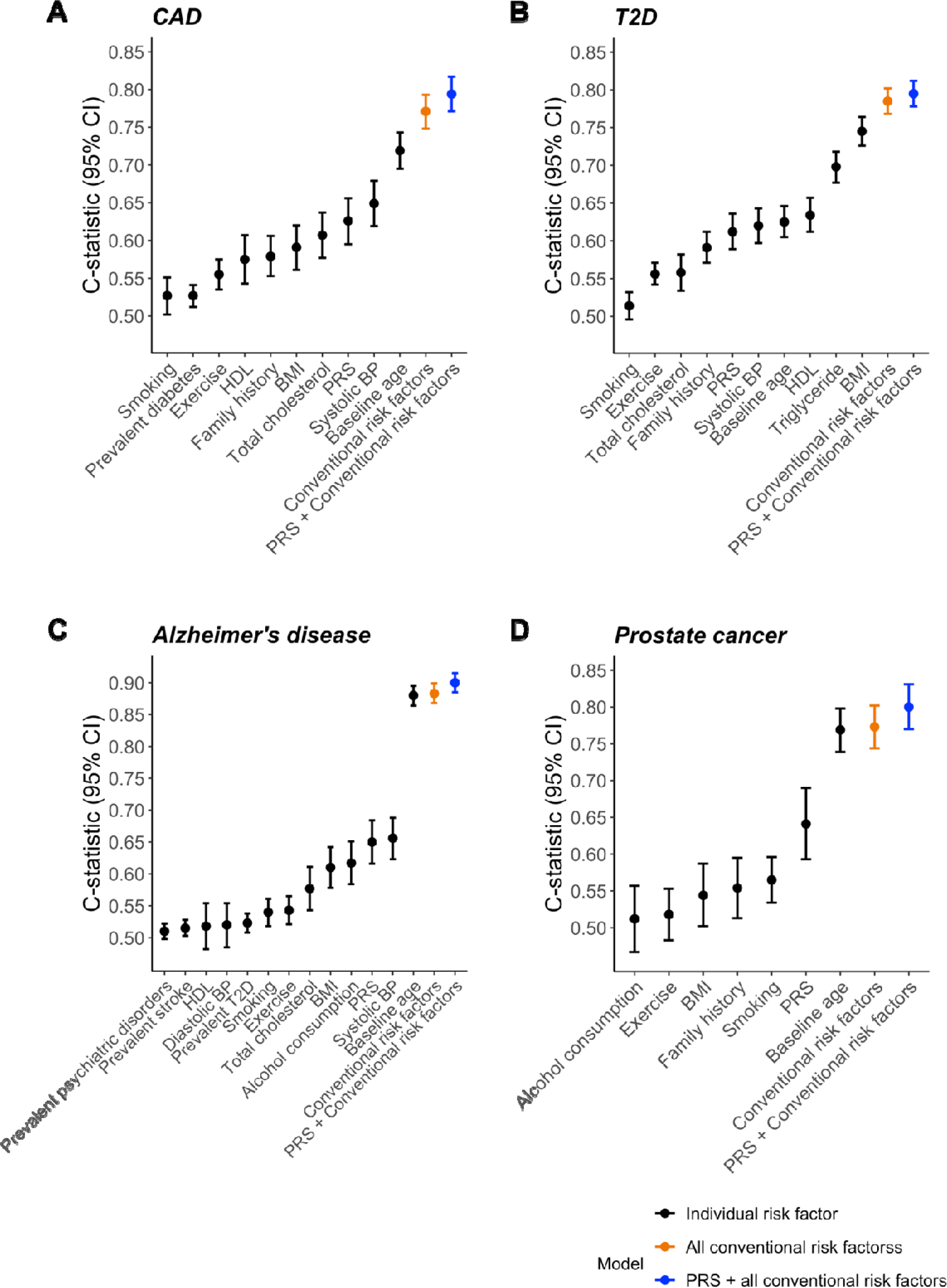
C-statistics of sex-stratified Cox models of disease-specific conventional risk factors and PRSs for incident (A) CAD, (B) T2D, (C) AD and (D) prostate cancer. C-statistics are depicted alongside their 95% confidence intervals. Models of individual risk factors are in black. Models combining disease-specific conventional risk factors are in orange. Models combining disease-specific conventional risk factors and PRSs are in blue.

In assessing the incremental gain in prediction of each PRS over its disease-specific conventional risk factors (**Figure 1**), we found ΔC-indices of 0.023 for CAD (95% CI 0.013-0.034), 0.01 for T2D (95% CI 0.004-0.016), 0.017 for AD (95% CI 0.010-0.024) and 0.027 for prostate cancer (95% CI 0.009-0.047). As expected, all PRSs were significantly associated with their respective incident diseases after adjusting for disease-specific conventional risk factors, and baseline age remained the strongest predictor for CAD, AD and prostate cancer (**Supplementary Figure 1**). We observed hazard ratios per standard deviation for PRS levels of 1.68 for CAD (95% confidence interval (CI) 1.50-1.88, P=2.25×10^-19^), 1.42 for T2D (95% CI 1.30-1.55, P=6.48×10^-15^), 1.92 for AD (95% CI 1.73-2.15, P=4.27×10^-32^), and 1.73 for prostate cancer (95% CI 1.47-2.04, P=5.50×10^-11^). The effects of PRS and family history were independent for incident CAD, T2D and prostate cancer, implying PRS and family history complement each other. As a sub-analysis for CAD, we excluded individuals taking anti-hypertensives and lipid-lowering medications at baseline (**Supplementary Figure 2A, B**) with the findings being consistent with the main analysis of all individuals.

For T2D, we performed a sub-analysis using NMR-determined glucose as an additional conventional risk factor (**Supplementary Figure 3A, B**). In sex-stratified Cox models of individual risk factors, BMI again had the strongest C-statistic (0.743, 95% CI 0.723-0.764), while PRS and glucose had C-statistics of 0.612 (95% CI 0.588-0.637) and 0.656 (95% CI 0.631-0.682), respectively. Adding PRS increased C-statistic over the model of conventional risk factors by 0.007 (95% CI 0.001-0.013). In the model combining PRS and conventional risk factors, PRS and glucose both were significantly associated with incident T2D with similar effect sizes (HR = 1.40 per s.d., 95% CI 1.27-1.54, P=1.85×10^-12^ and HR = 1.38 per s.d., 95% CI 1.28-1.48, P=5.95×10^-19^).

In a sub-analysis of AD in participants aged 60 and above (**Supplementary Figure 4**), the sex-stratified Cox model of PRS alone with C-statistic of 0.667 (95% CI 0.629-0.705) was greater than any individual conventional risk factor as well as the model combining all conventional factors. Adding PRS improved C-statistic over conventional risk factors by 0.064 (95% CI 0.036-0.096), leading to a model with C-statistic of 0.722 (95% CI 0.687-0.756). Notably, in the model combining PRS and all conventional risk factors of AD, PRS was associated with incident AD with an HR of 1.87 (95% CI 1.65-2.12, P=8.95×10^-23^) per s.d., which was greater than that for baseline age (HR=1.73 per s.d., 95% CI 1.51-1.98, P=4.50×10^-15^).

### Gut microbiome and incident disease

In FINRISK 2002, gut microbiome composition was determined by shallow shotgun metagenomic sequencing of baseline stool samples (**Methods**). To investigate the association between incident diseases and the overall variation in gut microbial communities, we performed Cox analyses on alpha and beta diversity at the species level adjusting for disease-specific conventional risk factors. Alpha diversity, estimated by Shannon index, was significantly negatively associated with incident T2D (HR 0.89 per s.d., 95% CI 0.82-0.96, P=0.004); no significant association was observed for incident CAD (HR 0.98 per s.d., 95% CI 0.88-1.09, P= 0.747), AD (HR 1.02 per s.d., 95% CI 0.90-1.15, P= 0.799) or prostate cancer (HR 1.09 per s.d., 95% CI 0.92-1.30, P= 0.325). In analysis of beta diversity using principal component analysis (PCA) of the Aitchison distance between taxonomic groups, incident T2D was associated with PC2 (HR 0.94, 95% CI 0.91-0.96, P= 1.31×10^-5^) and PC5 (HR 1.04, 95% CI 1.00-1.08, P=0.030).

To investigate the predictive capacity of gut microbial taxa for incident diseases, we focused on 235 species-level taxonomic groups after excluding rare and less prevalent taxa (**Methods**). In developing prediction models with taxa abundance at species levels, we utilised ridge logistic regression with 10×3-fold stratified cross validation (**Methods**). The average cross-validated area under the receiver operating characteristic curve (AUROC) of models were 0.597 (range 0.588-0.605) for CAD, 0.610 for T2D (0.599-0.624), 0.564 for AD (0.552-0.582) and 0.613 (0.595-0.626) for prostate cancer (**Supplementary Figure 5**). In sub-analyses, similar AUROC of cross-validated models were achieved for CAD (mean 0.587, range 0.552-0.609) and T2D (mean 0.604, range 0.589- 0.614), whereas the gut microbiome was not predictive of AD in participants aged 60 and above at baseline.

In sex-stratified (except for prostate cancer) Cox regression models, the gut microbiome score alone was significantly associated with all incident diseases (**Figure 2**), with HRs of 1.28 (95% CI 1.17- 1.41, P=2.29×10^-7^), 1.40 (95% CI 1.30-1.51, P=7.45×10^-20^), 1.34 (95% CI 1.20-1.50, P=2.09×10^-7^) and 1.50 (95% CI 1.27-1.78, P=1.66×10^-6^) per s.d. for incident CAD, T2D, AD and prostate cancer, respectively. After adjusting for disease-specific conventional risk factors (**Figure 2**), the effect of the gut microbiome score was significant but attenuated for incident T2D (HR = 1.20 per s.d., 95% CI 1.11-1.30, P=9.13×10^-6^) and prostate cancer (HR 1.23 per s.d., 95% CI 1.03-1.46, P=0.020); no significant effect of the gut microbiome score was found for CAD and AD. Compared with models of conventional risk factors, models adding the gut microbiome score yielded ΔC-statistics of 0.004 (95% CI 0-0.008) for T2D and 0.005 (95% CI -0.003-0.013) for prostate cancer. In the sub-analysis of T2D using NMR-based glucose as an additional conventional risk factor (**Supplementary Figure 3C**), the effect of the gut microbiome score was slightly attenuated (HR 1.16 per s.d., 95% CI 1.07-1.26, P=5.38×10^-4^) and the ΔC-statistic yielded by adding gut microbiome score to conventional risk factors was 0.003 (95% CI -0.001-0.006).

**Figure 2.**
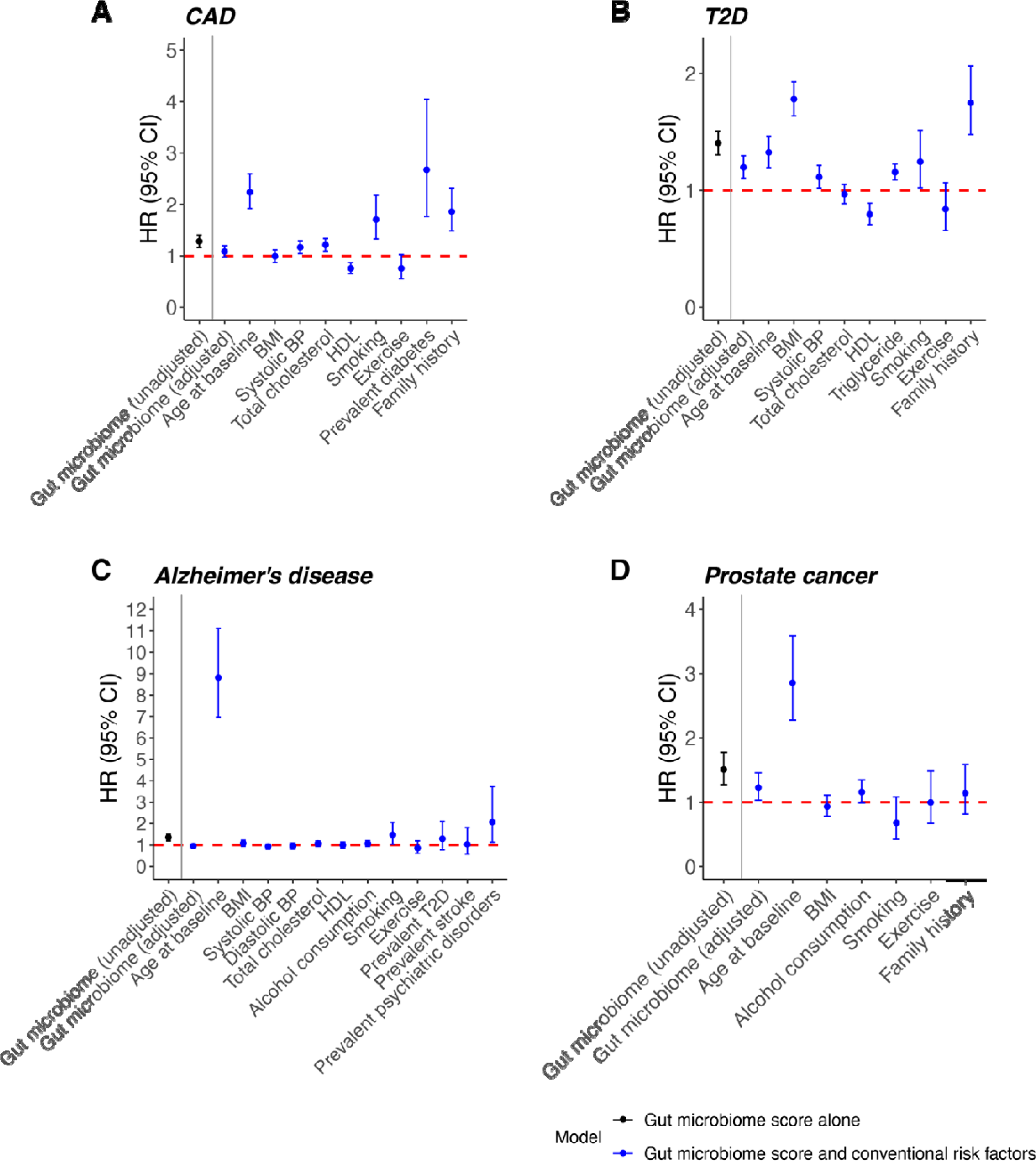
Hazard ratios (HR) and 95% confidence intervals (CI) for disease-specific gut microbiome scores and conventional risk factors for incident (A) CAD, (B) T2D, (C) AD and (D) prostate cancer. Sex-stratified Cox models of the gut microbiome score alone are in black and models combining the gut microbiome score and conventional risk factors are in blue.

### Integrative modelling of polygenic and gut metagenomic risk scores and conventional risk factors

We then investigated the combined predictive performance of PRS, the gut microbiome and conventional risk factors of their respective diseases using Cox regression models (**Table 2**). While age was the strongest individual predictor for incident CAD and prostate cancer, adding PRS and the gut microbiome score to age increased C-statistic by 0.049 (95% CI 0.030-0.066) and 0.032 (95% CI 0.011-0.052), respectively. For T2D, adding PRS and the gut microbiome score improved C-statistic over age by 0.076 (95% CI 0.057-0.095). For incident AD, adding PRS improved C-statistic over age by 0.019 (95% CI 0.011-0.026) while adding the gut microbiome score did not improve C-statistic. For all four diseases, the model combining disease-specific conventional risk factors, PRS and gut microbiome score achieved higher C-statistics than models based on any risk factors separately (**Table 2**). The combined model achieved ΔC-statistic over conventional risk factors of 0.024 (95% CI 0.013-0.035) for CAD, 0.014 (95% CI 0.007-0.021) for T2D, 0.017 (95% CI 0.009-0.024) for AD, and 0.031 (95% CI 0.011-0.05) for prostate cancer.

**Table 2.** C-statistics and 95% confidence intervals (CIs) of sex-stratified Cox regression models for PRS, gut microbiome score and conventional risk factors.

| Model<br>Disease | Age | Age+<br>PRS | Age+<br>microbiome<br>score | Age+PRS+<br>microbiome<br>score | Conventional<br>risk factors | Conventional<br>risk factors +<br>PRS<br>+ microbiome<br>score |
| --- | --- | --- | --- | --- | --- | --- |
|  | C-statistic (95% CI) |  |  |  |  |  |
| CAD | 0.719<br>(0.695-0.743) | 0.766<br>(0.742-0.789) | 0.722<br>(0.698-0.747) | 0.767<br>(0.744-0.791) | 0.771<br>(0.748-0.793) | 0.794<br>(0.772-0.817) |
| T2D | 0.625<br>(0.605-0.646) | 0.675<br>(0.654-0.695) | 0.665<br>(0.644-0.685) | 0.702<br>(0.681-0.722) | 0.785<br>(0.768-0.802) | 0.799<br>(0.783-0.816) |
| AD | 0.880<br>(0.864-0.895) | 0.898<br>(0.883-0.914) | 0.880<br>(0.864-0.895) | 0.898<br>(0.883-0.914) | 0.883<br>(0.868-0.899) | 0.900<br>(0.885-0.915) |
| Prostate<br>cancer | 0.769<br>(0.739-0.798) | 0.797<br>(0.766-0.828) | 0.774<br>(0.745-0.802) | 0.801<br>(0.770-0.832) | 0.773<br>(0.744-0.802) | 0.804<br>(0.774-0.834) |

The sub-group analyses for CAD, T2D and AD showed consistent results in general. In a sex-stratified Cox model for CAD (**Supplementary Figure 2D**), adding PRS and the gut microbiome score increased C-statistic by 0.050 (95% CI 0.030-0.068) over age, and 0.025 (95% CI 0.013-0.038) over all conventional risk factors in individuals without baseline use of anti-hypertensives or lipid-lowering medications. For T2D (**Supplementary Figure 3D**), adding PRS and gut microbiome score improved C-statistic over age by 0.073 (0.051-0.092), and the combined model increased C-statistic by 0.010 (95% CI 0.003-0.016) as compared to the model of conventional risk factors including NMR-based glucose. In the sub-group analysis for AD in those older than 60 years at baseline, adding PRS improved C-statistic over baseline age by 0.077 (95% CI 0.043-0.108), while the gut microbiome did not show improvement.

In the combined models (**Supplementary Tables 1-4)**, PRSs were found to be significantly associated with CAD (HR per s.d. 1.68, 95% CI 1.50-1.88, P=4.39×10^-19^), T2D (HR per s.d. 1.41, 95% CI 1.29- 1.54, P=1.38×10^-14^), AD (HR per s.d. 1.93, 95% CI 1.73-2.15, P=3.85×10^-32^) and prostate cancer (HR per s.d. 1.72, 95% CI 1.46-2.02, P=1.05×10^-10^). The gut microbiome score was associated with T2D (HR per s.d. 1.19, 95% CI 1.10-1.29, P=2.11×10^-5^) and prostate cancer (HR per s.d. 1.19, 95% CI 1.01-1.41, P=0.041).

In subgroup analyses (**Supplementary Tables 5-7**), similar effects of PRSs were found for CAD (HR per s.d. 1.77, 95% CI 1.56-2.02, P=3.05×10^-18^), T2D (HR per s.d. 1.40, 95% CI 1.27-1.53, P=3.43×10^-12^) and AD (HR per s.d. 1.88, 1.65-2.13, P=8.33×10^-23^); the effect of the gut microbiome score remained significant for T2D (HR per s.d. 1.15, 95% CI 1.06-1.25, P=1.07×10^-3^) after adjusting for NMR-based glucose and other conventional risk factors.

## Discussion

While the interplay between host genetics and gut microbiome has been increasingly recognised and studied[30, 44, 45], few studies have investigated their combined impact on complex disease risk. This study presents a joint analysis of genotyping data, gut metagenomics data and clinical metadata for four common complex diseases (CAD, T2D, AD and prostate cancer) in a large prospective population-based cohort. We compared popular published PRSs for each disease, baseline metagenomics of gut microbiota, and conventional risk factors for predicting onset of each disease over a median of 17.8 years of follow-up. We demonstrated that PRS improved prediction performance over conventional risk factors for all diseases studied. However, unlike PRSs, while the gut microbiome improved prediction over age for CAD, T2D and prostate cancer, there was little evidence that gut microbiome improved prediction performance when modelled jointly with conventional risk factors. The information (e.g. features and coefficients) necessary to independently apply our integrated predictive models are provided in **Supplementary Tables 1-4**.

As expected, in our study higher PRS was significantly associated with higher disease incidence for all four diseases, consistent with previous studies. Also expected, we found PRS for all four diseases improved predictive ability over conventional risk factors, adding to the body of evidence[9, 14] that PRS have potential clinical utility complementing traditional risk factors. Consistent with prior work, we demonstrated that PRSs improved prediction of CAD, T2D and prostate cancer independently of and in addition to family history, a strong risk factor for all diseases studied [46–50]. Notably, for AD, whose risk of development attributed to genetics has been estimated at 70%[51], the PRS improved C-statistic over conventional risk factors including age by 0.017 in all studied participants and 0.064 in participants aged 60 and above at baseline.

Although the ΔC-statistics for gut microbiome scores over conventional risk factors were small, we observed significant improvement in sex-stratified prediction models over baseline age alone for CAD, T2D and prostate cancer [26, 52–54]. In accordance with previous studies, we found a significant inverse signal between baseline alpha diversity and incident T2D[55], which might be partially explained by possible mediation effects of gut microbiota-derived metabolites correlating with lower microbial diversity (e.g., imidazole propionate) and insulin resistance[56, 57]. We also found significant associations between beta diversity and incident T2D, which might indicate a shift in microbiome composition involved in disease pathogenesis and progression[26, 58, 59]. The results suggest that the physiological and metabolic processes influenced by risk-associated changes in gut microbiome are largely captured by conventional risk factors for the diseases studied.

Our study has limitations. First, the gut microbiome and conventional risk factors were measured only once at the initial assessment. Although the gut microbiome remains largely stable during adulthood, the microbial community is influenced by environment and cohabitation in the long term[60–62], thus their effects on future disease may change from what we estimate here. Second, due to unavailability, we did not assess the impact of family history of Alzheimer’s disease, a risk factor that may also capture important aspects of shared environment influencing gut microbiome composition[63, 64]. Third, the generalisability of the microbiome and integrated risk models to other external cohorts could not be investigated due to the paucity of large prospective studies with similar data types. The composition of the human gut microbiome differs across geographically and culturally distinct settings, which can be attributed to variations in host genetics, immunity, and behavioural features[65, 66]. Lastly, our study cohort comprised European ancestry (Finnish) participants, thus predictive performance of PRS and improvement over conventional risk factors may not generalize to other demographics and healthcare systems, particularly as the predictive performance of PRS derived in Europeans are known to be attenuated when applied to populations of non-European ancestries[67–69].

In summary, this work presents one of the first studies on prediction of incident common complex diseases integrating PRSs, gut metagenomics and clinical metadata. Our study highlights potential limitations in the use of the human gut microbiome for improving clinical risk prediction despite its association with incident disease; however, larger studies are warranted in order to better quantify potential incremental gains. Overall, we show that integrating PRS and gut metagenomic scores can maximise predictive capacity for common diseases over conventional risk factors alone.

## Materials and Methods

### Study design

The FINRISK surveys have been conducted to investigate risk factors for major chronic non-communicable diseases every 5 years since 1972 in Finland[70]. This work was based on FINRISK study carried out in 2002. The study included independent and representative population samples of six geographical areas of Finland: (1) North Karelia, (2) Northern Savo, (3) Turku and Loimaa, (4) Helsinki and Vantaa, (5) Oulu and (6) Lapland, that were randomly drawn from the National Population Information System[42]. With an overall participant rate of 65%, the FINRISK 2002 cohort comprised a total of 8,783 individuals out of 13,498 invitees aged 25 to 74 years. The participants filled in self-administered questionnaires, undertook health examinations conducted by trained personnel at the study sites, and donated biological samples including venous blood and stool. All participants gave written informed consent, and the study protocol was approved by the Coordinating Ethics Committee of the Helsinki University Hospital District (Ref. 558/E3/2001). The surveys were conducted in accordance with the World Medical Association’s Declaration of Helsinki on ethical principles. In the present study, we included individuals whose genotyping data and shotgun metagenomics sequencing of stool samples were both available. We excluded individuals with (1) low reads of metagenomic sequencing (total mapped reads <100,000), (2) baseline pregnancy, (3) BMI>=40 kg/m^2^ or <16.5 kg/m^2^ and (4) antibiotic use up to one month prior to baseline. Altogether, samples from 5,676 participants were eligible for this study.

### Baseline examination and sample collection

Demographic factors, physiological measurements, lifestyle factors, biomarkers and biological samples were collected at baseline in 2002[42]. Questionnaires and invitation to health examinations were mailed to all subjects. Self-administered questionnaires included information such as participant’s background, medical history, diet, and self-reported family history of some diseases. Questionnaires were in paper form and saved to electrical format. The health examination and blood sampling were performed by trained nurses at local health centres or other survey sites. Physical measurements such as weight, height and blood pressure were obtained during the health examination. Venous blood samples were collected for the full cohort. The samples were collected after the participants were fasted for ≥4ℒh and centrifuged at the field survey sites. The fresh samples were transferred daily to the central laboratory of the Finnish Institute for Health and Welfare and analyzed during the two following days.

Stool samples were collected from willing participants by using stool sampling kits with detailed instructions at home. The samples were mailed overnight to the laboratory of the Finnish Institute for Health and Welfare under winter conditions in Finland and immediately stored at -20°C upon receipt. The stool samples were kept unthawed until 2017 when they were transferred to the University of California San Diego for sequencing.

### Disease endpoints, exclusion criteria and factors

We studied four incident diseases: coronary artery disease (CAD), type 2 diabetes (T2D), Alzheimer’s disease (AD) and prostate cancer. The participants were followed up until December 31^st^, 2019 using electronic health records (EHR) linkage to the Finnish national registries. Disease cases were identified based on International Classification of Diseases (ICD) codes, Anatomical Therapeutic Chemical (ATC) codes, from the Care Register for Health Care (hospital discharges and specialized outpatient care), Finnish Cancer Register and the Drug Reimbursement and Purchase Registers. CAD cases were defined by ICD-10 I20.0|I21|I22, ICD-9 410|4110, ICD-8 410|4110; T2D cases were defined by ICD-10 E1[1–4], ICD-9 250, ICD-8 250, KELA drug reimbursement code 215 and ATC A10B; Alzheimer’s disease cases were defined by ICD-10 G30|F00, ICD-9 331.0, ICD-8 290.10, KELA reimbursement code 307, reimbursement with ICD code G30|F00|3110 and ATC N06D; prostate cancer cases were identified in the Finnish Cancer Register. Follow-up time was extracted from EHRs and determined by the years to the first incident event, or death, or end of the follow-up study period.

The conventional risk factors for CAD were defined as follows: age, sex, body mass index (BMI), systolic blood pressure (SBP), total cholesterol, high-density lipoprotein (HDL) cholesterol, current smoking status, exercise, any prevalent diabetes and parental history of myocardial infarction[12]. Smoking status was defined as current use of tobacco products at baseline. Exercise was defined as regular exercise for at least 3 hours/week or regular competitive sports training according to responses to self-administered questionnaires. Individuals with missing values of risk factors were excluded. Individuals with prevalent diagnosis of heart diseases were excluded. A total of 5,093 individuals were considered for CAD analyses. In the sub-analysis of CAD, participants with baseline use of antihypertensives or lipid-lowering medications were further excluded, resulting in a subset of 4,293 individuals.

For T2D, the risk factors included age, sex, BMI, SBP, total cholesterol, HDL, triglycerides, current smoking status, exercise and parental history of any diabetes[26, 47]. After individuals with incomplete values of risk factors, any prevalent diabetes, baseline use of diabetes medication, and glycated haemoglobin (HbA1c) (if available) >= 6.5% were excluded, a total of 5,297 individuals were involved in T2D analyses. In an additional sub-analysis of T2D, baseline glucose determined by the Nightingale Health NMR platform from frozen serum samples was included as an additional risk factor in a subset of 4,911 individuals.

For Alzheimer’s disease, the risk factors included age, sex, BMI, SBP, diastolic blood pressure (DBP), total cholesterol, HDL, average weekly alcohol consumption, current smoking status, exercise, prevalent T2D, prevalent stroke and any prevalent psychiatric disorders including depression, bipolar disorder and schizophrenia[71]. We excluded individuals with missing values of risk factors and prevalent dementia, which resulted in 5,347 individuals for analyses of Alzheimer’s disease. The sub-analysis of AD in participants aged 60 and above at baseline included 1,220 individuals.

For prostate cancer analyses, the risk factors included age, BMI, average weekly alcohol consumption, exercise, current smoking status and parental history of any cancer[72]. Only male participants were studied. After individuals with incomplete risk factors and prevalent diagnosis of prostate cancer were excluded, a total of 2,464 individuals remained for analyses of prostate cancer.

### Characterization of gut microbiome

The library generation was carried out with a miniaturised version of the Kapa HyperPlus Illumina-compatible library prep kit (Kapa Biosystems)[73]. The DNA extracts were normalized to 5ℒng total input per sample using an Echo 550 acoustic liquid-handling robot (Labcyte Inc). Enzymatic fragmentation (1/10 scale), end-repair and adapter-ligation reactions were performed using a Mosquito HV liquid-handling robot (TTP Labtech Inc.). Sequencing adapters were based on the iTru protocol[74]. Amplified and barcoded libraries were quantified by the PicoGreen assay and sequenced on an Illumina HiSeq 4000 instrument to an average depth of ∼900,000 reads/sample. The stool shotgun sequencing was successfully performed in 7,231 individuals. The shotgun metagenomic sequences were analyzed with an automated Snakemake workflow pipeline (https://github.com/tanaes/snakemake_assemble)[73, 75]. Adapters and low-quality sequences were trimmed with Atropos[76], and host reads were removed with Bowtie2[77] against the human genome assembly GRCh38.

Stool metagenomes were classified using Kraken2[78] and a custom index database based on species definitions from 258,406 reference genomes from GTDB release R06-RS202 (27th April 2021)[79]. Bracken[80] was used to re-estimate abundances after Kraken2 classification. A threshold of 250 reads/taxon was used to define a positive hit which resulted in 4026 species identified with a mean prevalence rate of 4.74%. After removing samples with total mapped read counts lower than 100,000 reads/sample, taxonomic profiles from 7,205 individuals were retained for analyses.

### Genotype data processing and polygenic score calculation

Genotyping was undertaken using Illumina genome-wide SNP arrays (the HumanCoreExome BeadChip, the Human610-Quad BeadChip and the HumanOmniExpress)[49]. After samples with ambiguous gender, missingness > 5%, excess heterozygosity and non-European ancestries were removed, and variants with missingness > 2%, Hardy-Weinberg equilibrium P value < 1×10^-6 and minor allele count < 3 were excluded, the samples were pre-phased with Eagle2 v2.3. A Finnish- population-specific reference panel consisting of 2,690 high-coverage whole-genome sequencing and 5,092 whole-exome sequencing samples was used with IMPUTE2 v2.3.2 to perform genotype imputation. Post-imputation quality control was applied using PLINK v.2.0. Variants with INFO score < 0.7, minor allele frequency < 1%, Hardy–Weinberg equilibrium P < 1×10^-6 were excluded. Samples with missing rate > 10% were excluded. A total of 7,967,866 variants and 7,281 samples remained after quality control.

For all diseases studied, we calculated polygenic risk scores (PRSs) in FINRISK 2002 cohort using external summary statistics in the Polygenic Score Catalog[43]. We considered previously published scores that were developed mainly based on large European populations and did not include FINIRSK 2002 participants in their development The Polygenic Score Catalog IDs of the PRSs for CAD, T2D, AD and prostate cancer were PGS000018[12], PGS000036[37], PGS000334[38] and PGS000662[40], respectively. Each PRS was computed by multiplying the genotype dosage of each risk allele at each variant by its weight and summing across all variants in the respective score with PRSice-2[81]. The final PRSs consisted of 1,396,966 variants for the CAD PRS, 129,793 for the T2D PRS, 21 for the AD PRS, and 181 for the prostate cancer PRS.

### Statistical analysis

Statistical analysis was performed with R versions 4.2.1 and 3.6.0. Cox proportional hazards models stratified by sex were first fit for time-on-study for each incident disease on each of their respective conventional risk factors and PRS separately. Next, a model combining disease-specific PRS and conventional risk factors was fit for each disease. Prostate cancer was studied only in men; its respective analysis did not include sex stratification. The ability of models to distinguish between cases and non-cases were assessed and compared with the Harrell’s C-statistic, a performance metric for evaluating model discrimination based on censored survival data. Proportional hazards assumptions were examined by Schoenfeld residuals. Hazard ratios (HR), 95% confidence intervals (CI), and two-sided Wald test Pℒvalues were reported for risk factors. Statistical significance was determined with P-value threshold of 0.05.

The gut microbiota diversities were measured with species-level abundance data before filtering taxa by relative abundance and prevalence. Alpha diversity of the gut microbiome was calculated with Shannon index using raw counts. Rarefaction was not performed to avoid loss of data and samples had total mapped reads >100,000 after filtering. Beta diversity was estimated separately in samples by applying principal component analysis (PCA) on centered log-ratio (CLR) transformed abundance data, i.e., using the Aitchison distance, after disease-specific exclusion criteria were applied. Cox proportional hazards models were fit for time-on-study for each disease on gut microbiome alpha diversity and the first 5 principal components of CLR abundance adjusting for conventional risk factors and stratified by sex (excepting prostate cancer analyses).

We subsequently focused on common and abundant taxa that were detected with prevalence >1% and relative abundance of >0.1% in at least 10% of samples. After excluding rare and less prevalent taxa, 235 species-level taxonomic groups were obtained and CLR-transformed for prediction modelling. For each incident disease studied, we evaluated the predictive capacity of the gut microbiome composition using Ridge logistic regression models of species-level CLR abundance with repeated cross-validation (3-fold, repeated 10 times) stratified for disease status. The optimal lambda value was determined from a grid search space ranging from 0.0001 to 100. The prediction performance was assessed using area under the receiver operating characteristic curve (AUC). In assessing the association between the gut microbiome risk score and incident disease, we used the predicted scores based on the optimal cross-validated models of the gut microbiome composition. For each disease studied, sex-stratified (except for prostate cancer) Cox regression model was fit for time-on-study on the gut microbiome score by itself and with adjustment of disease-specific conventional risk factors.

Finally, we investigated whether disease-specific PRSs and microbiome scores made independent contributions to predicting disease risk. For each incident disease, sex-stratified (expect for prostate cancer) Cox models were fit on disease-specific PRSs and microbiome scores separately and in combination, adjusting for age at baseline; cox models were also fit on baseline age alone for comparison. Sex-stratified (expect for prostate cancer) Cox models were then fit on disease-specific PRS, gut microbiome score and conventional risk factors, and compared to Cox models combining disease-specific conventional risk factors. Covariates and their respective coefficients in Cox regression models for all diseases studied were reported in **Supplementary Tables 1-4**.

### Data availability

The FINRISK data for the present study are available with a written application to the THL Biobank as instructed on the website of the Biobank (https://thl.fi/en/web/thl-biobank/for-researchers). A separate permission is needed from FINDATA (https://www.findata.fi/en/) for use of the EHR data. Metagenome data are available in European Genome-Phenome Archive via EGAD00001007035.

## Acknowledgements

YL was supported by funding from the Cambridge Baker Centre for Systems Genomics. SR was supported by a British Health Foundation programme grant (RG/18/13/33946). MOR was funded by the Research Council of Finland (grant no. 338818). LL was supported by the European Union’s Horizon 2020 research and innovation program (grant no. 952914). TN was supported by the Finnish Foundation for Cardiovascular Research, the Sigrid Jusélius Foundation, the Southwestern Finland Hospital District, and the Research Council of Finland (grants no. 321351 and 354447). VS was supported by the Finnish Foundation for Cardiovascular Research and by the Juho Vainio Foundation. ASH was supported by the Research Council of Finland, grant no. 321356. MI was supported by the Munz Chair of Cardiovascular Prediction and Prevention and the NIHR Cambridge Biomedical Research Centre (BRC-1215-20014; NIHR203312) [*]. MI was also supported by the UK Economic and Social Research 878 Council (ES/T013192/1). This study was supported by the Victorian Government’s Operational Infrastructure Support (OIS) program and by core funding from the British Heart Foundation (RG/18/13/33946) and the NIHR Cambridge Biomedical Research Centre (BRC- 1215-20014; NIHR203312) [*]. *The views expressed are those of the author(s) and not necessarily those of the NIHR or the Department of Health and Social Care. This work was supported by Health Data Research UK, which is funded by the UK Medical Research Council, Engineering and Physical Sciences Research Council, Economic and Social Research Council, Department of Health and Social Care (England), Chief Scientist Office of the Scottish Government Health and Social Care Directorates, Health and Social Care Research and Development Division (Welsh Government), Public Health Agency (Northern Ireland), British Heart Foundation and Wellcome.

