## Supplementary Figure for "Integration of polygenic and gut metagenomic risk prediction for common diseases"

**Supplementary Figure 1.** Sex-stratified Cox models of disease-specific conventional risk factors and PRSs showing significant associations between PRS and incident diseases for (A) CAD, (B) T2D, (C) AD and (D) prostate cancer. HRs of risk factors are depicted alongside their 95% confidence intervals.

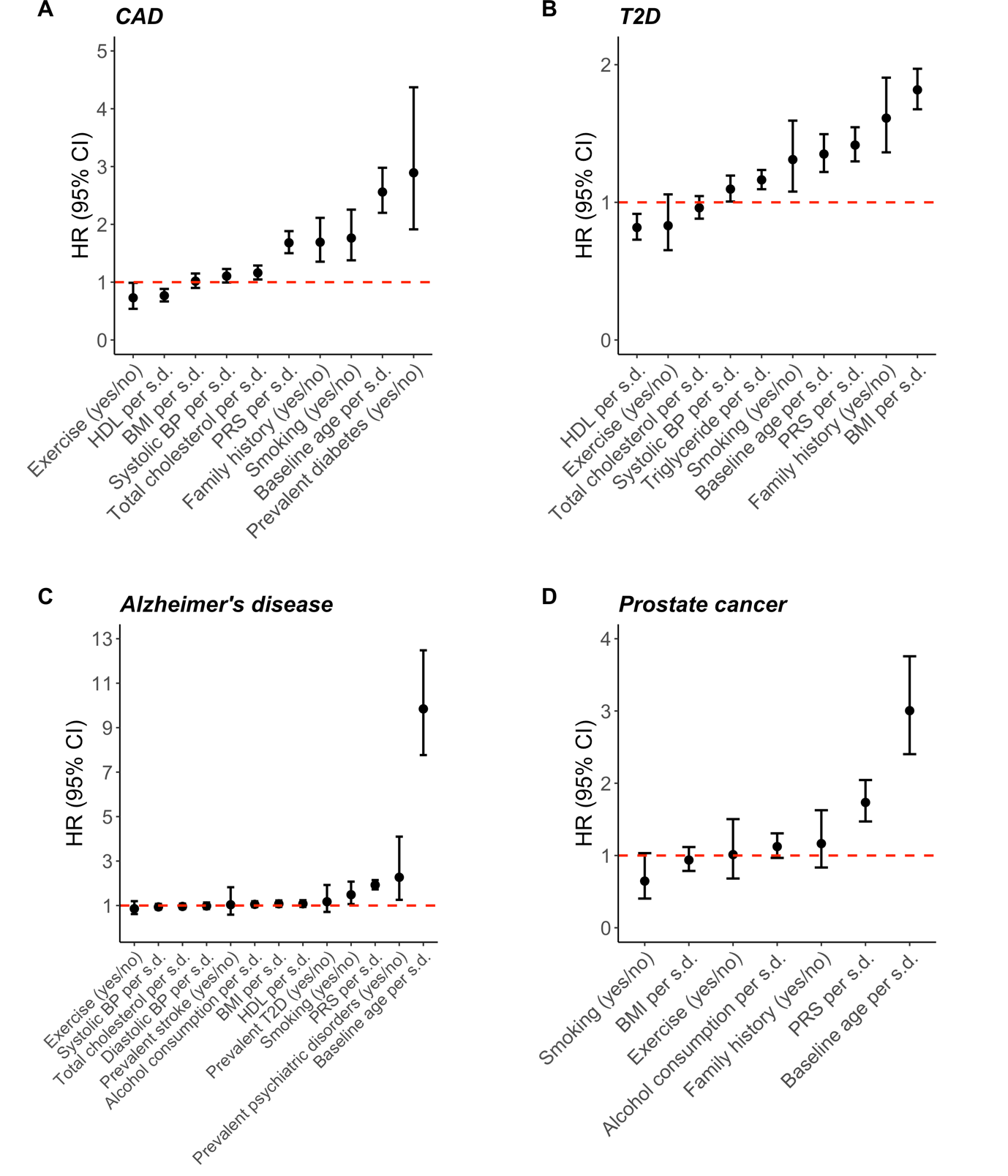

**Supplementary Figure 2.** Sub-analysis of incident CAD in individuals who were not on antihypertensives and lipid-lowering medications at baseline. In sex-stratified Cox models of conventional risk factors and PRS, (A) C-statistics and (B) HRs are depicted alongside their 95% confidence intervals; models of individual factors are in black; the model combining conventional risk factors is in orange; the model combining conventional risk factors and PRS is in blue. In sex-stratified Cox models of conventional risk factors and the gut microbiome score, (C) HRs of the gut microbiome score and conventional risk factors are depicted alongside their 95% confidence intervals; the model of the gut microbiome score alone is in black; the model combining the gut microbiome score and conventional risk factors is in blue. (D) The C-statistics and 95% confidence intervals of Cox models for integrative analysis.

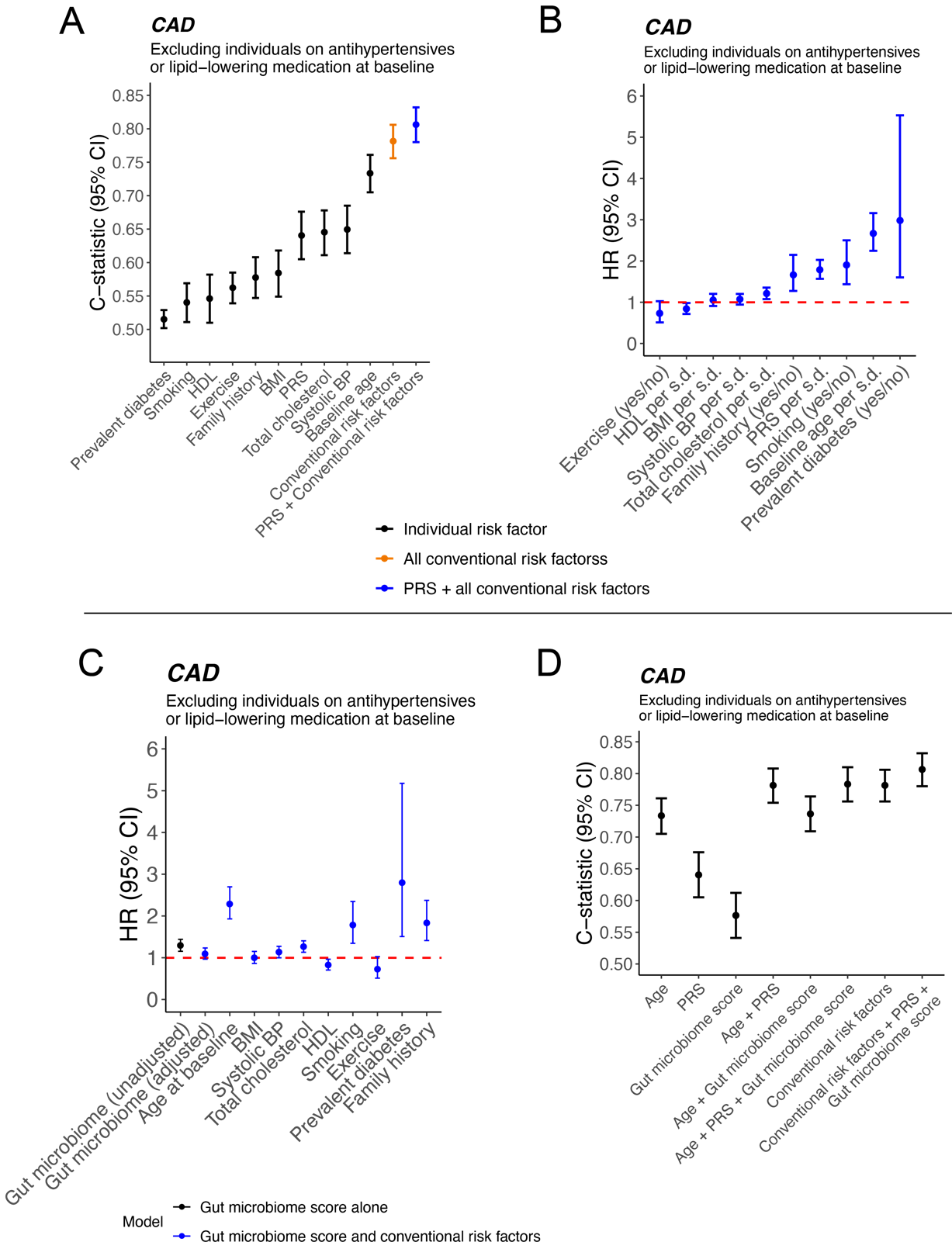

**Supplementary Figure 3.** Sub-analysis of incident T2D using NMR-determined glucose as an additional risk factor in sex-stratified Cox models. In Cox models of conventional risk factors and PRS, (A) C-statistics and (B) HRs are depicted alongside their 95% confidence intervals; models of individual risk factors are in black; the model combining conventional risk factors is in orange; the model combining conventional risk factors and PRSs is in blue. In sex-stratified Cox models of conventional risk factors and the gut microbiome score, (C) HRs of the gut microbiome score and conventional risk factors are depicted alongside their 95% confidence intervals; the model of the gut microbiome score alone is in black; the model combining the gut microbiome score and conventional risk factors is in blue. (D) The C-statistics and 95% confidence intervals of Cox models for integrative analysis.

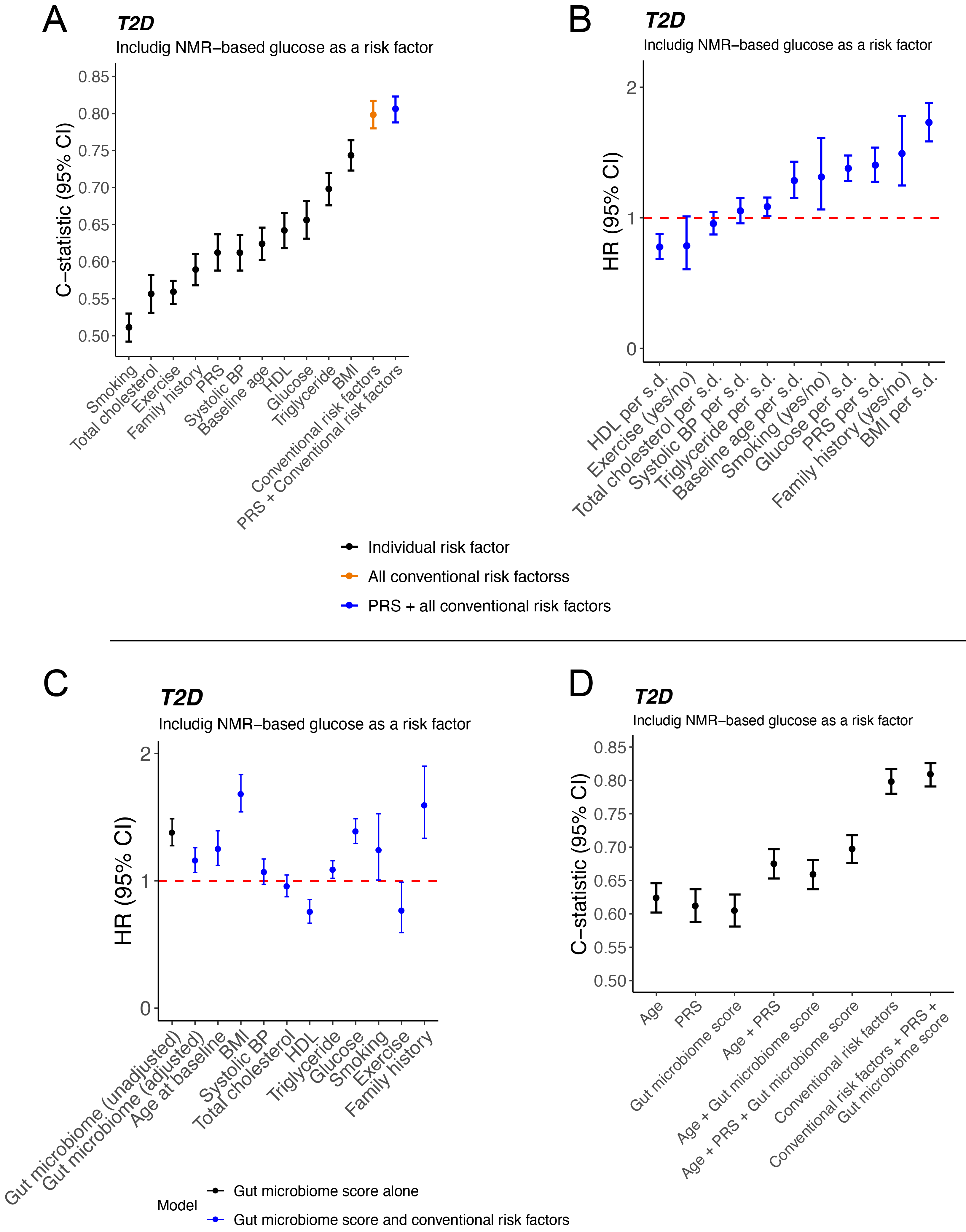

**Supplementary Figure 4.** Sub-analysis of incident AD in participants aged 60 and above at baseline using sex-stratified Cox models. (A) C-statistics and (B) HRs of conventional risk factors and PRS are depicted alongside their 95% confidence intervals. Models of individual risk factors are in black. Models combining disease-specific conventional risk factors are in orange. Models combining disease-specific conventional risk factors and PRSs are in blue.

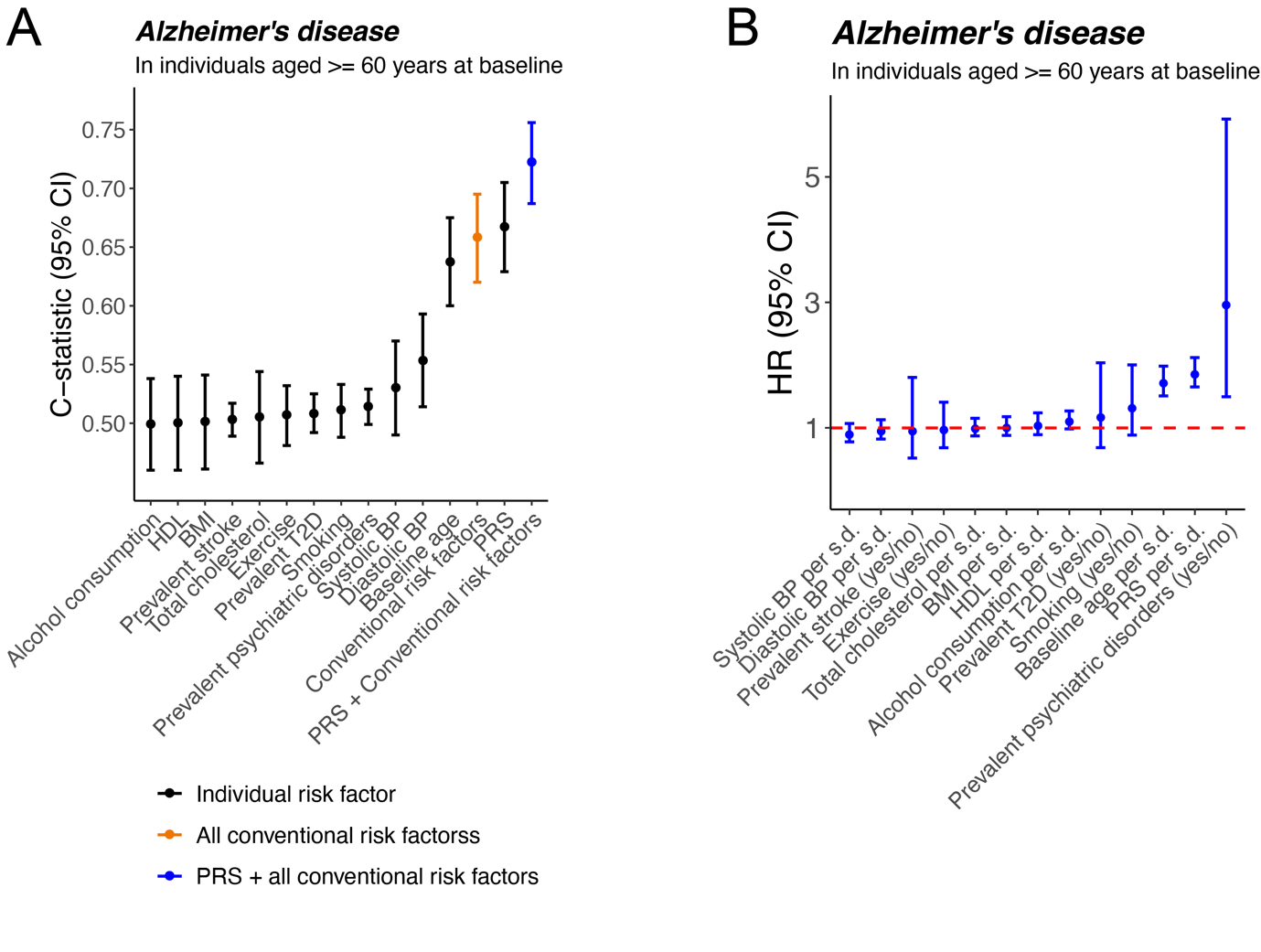

**Supplementary Figure 5.** Cross-validated Ridge logistic regression models for incident (A) CAD, (B) T2D, (C) AD and (D) prostate cancer using gut microbiome composition. The ROC curve of the optimal cross-validated model is in red and curves of other models are in grey.

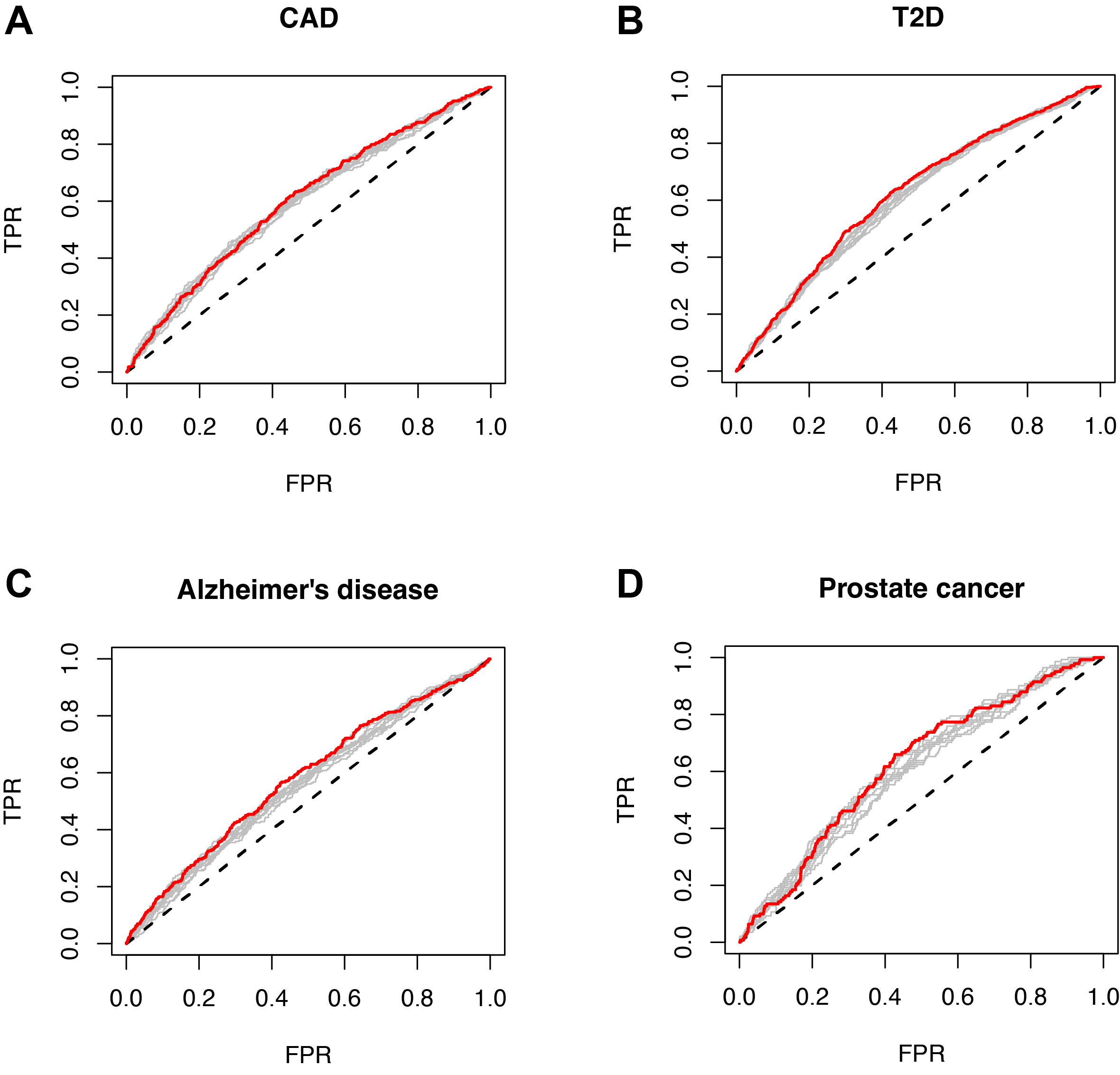
